# Prevalence and uptake of vaping among people who have quit smoking: a population study in England, 2013-2024

**DOI:** 10.1101/2024.07.23.24310879

**Authors:** Sarah E. Jackson, Jamie Brown, Loren Kock, Lion Shahab

**Author notes:** Corresponding author: Dr Sarah Jackson, Department of Behavioural Science and Health, University College London, 1-19 Torrington Place, London WC1E 7HB, UK.

## Abstract

**Background:** Vaping prevalence has increased rapidly in England since 2021. This study estimated trends between 2013 and 2024 in vaping among ex-smokers, overall and among those who did not use e-cigarettes to support their quit attempt.

**Methods:** Data were collected via nationally-representative, monthly cross-sectional surveys in England, October-2013 to May-2024. We analysed data from 54,251 adults (≥18y) who reported having tried to stop smoking in the past year or having stopped smoking more than a year ago. Logistic regression estimated associations between time and e-cigarette use.

**Findings:** Across the period, there were increases in the use of e-cigarettes to support attempts to stop smoking (from 26.9% [24.0-30.0%] in October-2013 to 41.4% [37.7-45.2%] in May-2024), in current vaping among ≥1y ex-smokers (1.9% [1.5-2.5%] to 20.4% [18.7-22.2%]), and in late uptake of vaping after smoking cessation (i.e., current vaping among people who quit smoking before e-cigarettes started to become popular in 2011; 0.4% [0.2-0.8%] to 3.7% [2.8-4.9%]). These increases were non-linear, with much of the change occurring since mid-2021, and were greatest at younger ages (e.g., current vaping among ≥1y ex-smokers reached 58.9% among 18-year-olds vs. 10.7% among 65-year-olds).

**Interpretation:** Vaping prevalence increased substantially among adult ex-smokers in England over the past decade, particularly at younger ages. While this is likely to have been largely driven by increased use of e-cigarettes in quit attempts and continued use thereafter, there was also evidence of increased uptake of vaping among those who had been abstinent from smoking for many years.

**Funding:** Cancer Research UK.

**Research in context:** *Evidence before this study:* We searched PubMed up to July 2024 using the following search terms (“e-cigarette∗” OR “vap∗”) AND (“trend*”) AND (“ex-smok*” OR “former smok*” OR “quit smok*” OR “smoking status”) AND (“adult*”) AND (“England” OR “Great Britain” OR “United Kingdom”). The search returned 13 results, of which four reported on changes in the prevalence of current vaping among adults who have stopped smoked. The first documented an increase in current vaping among >1y ex-smokers between 2010 and 2013 (from 0.3% to 2.7%). The second indicated the prevalence of current vaping among ≥1y ex-smokers rose further in subsequent years (from 3.3% in 2014 to 10.4% in 2019) and also reported an increase among people who quit smoking before e-cigarettes started to become popular in 2011 (from 0.8% in 2014 to 2.1% in 2019). The third reported a substantial increase in the proportion of ≥1y ex-smokers who were vaping with disposable e-cigarettes between 2021 and 2023 (from 0.0% in January 2021 to 4.1% in August 2023). The fourth reported an increase in long-term (>6 month) vaping among ≥1y ex-smokers between 2013 and 2023 (from 1.4% in October 2013 to 16.2% in October 2023), with much of this increase occurring since 2021.

*Added value of this study:* This study shows the proportion of adult ex-smokers in England who vape increased substantially between 2013 and 2024. As of May 2024, there were around 2.2 million adult vapers in England who had stopped smoking. There were also increases in the use of e-cigarettes in attempts to quit smoking and in the uptake of vaping among people among people who quit smoking before e-cigarettes started to become popular. These increases were non-linear, with much of the change occurring since mid-2021, and were greatest at younger ages

*Implications of all the available evidence:* Since disposable e-cigarettes were introduced to the market in 2021, there has been a sharp increase in vaping among ex-smokers. While this is likely to have been largely driven by increases in people using e-cigarettes as a smoking cessation aid and continuing to vape beyond their successful quit attempt, there was also evidence of increased uptake of vaping among those who had been abstinent from smoking for many years.

## Introduction

There is good evidence from randomised controlled trials^1^ and real-world studies^2,3^ that e-cigarettes help people to quit smoking. E-cigarette use is behaviourally similar to cigarette smoking and the devices deliver nicotine effectively.^4^ They are generally regarded as much less harmful than combustible tobacco, but pose some risks compared with neither smoking nor vaping.^5^ The extent to which vaping protects against or increases the risk of relapse to smoking in the longer term is not yet clear. Given ex-smokers represent a growing proportion of the adult population,^6^ monitoring patterns of vaping among people who have quit smoking is important because unless vaping prevents relapse to smoking, it will expose users to some level of additional harm.^5^

E-cigarettes were first introduced to the UK market in 2008. They were rarely used up to 2011,^7^ then rapidly became popular as a method of quitting smoking. Up to mid-2011, fewer than one in 100 quit attempts in England involved the use of an e-cigarette; by 2014 this number had risen to more than one in four.^8^ In the years that followed, the proportion of ex-smokers who had been quit for ≥1 year who reported current vaping increased steadily, from 3.3% in 2014 to 10.4% in 2019.^9^ While most ex-smokers who vape likely start using e-cigarettes while smoking and continue to vape beyond a successful quit attempt, some appear to take up vaping *after* stopping smoking. Between 2014 and 2019, 7.1% of ex-smokers who had been quit for <1 year who did not use an e-cigarette in their quit attempt reported current vaping, as did an increasing proportion of ex-smokers who quit before e-cigarettes became popular in 2011 (from 0.8% in 2014 to 2.1% in 2019).^9^

Since 2021, there has been a substantial increase in vaping in England among all smoking statuses, which appears to have been linked to the introduction of new disposable e-cigarettes.^10,11^ The proportion of ≥1y ex-smokers who reported having vaped for more than 6 months doubled between the start of 2021 and October 2023 (from 8% to 16%)^10^ and the proportion currently using disposable e-cigarettes increased from 0% to 4%.^11^ Studies show the recent increase in vaping has been much greater among younger adults and those who drink more heavily.^10–12^ It is not clear whether the same patterns have occurred among ex-smokers specifically, or whether there have been differences by other key sociodemographic characteristics (e.g., gender or socioeconomic position). It is also not clear to what extent the increase in vaping prevalence among ex-smokers reflects more people taking up vaping after smoking cessation or a change in the types of ex-smokers who are vaping.

This study aimed to understand more about vaping among adults in England who have quit smoking, including the extent to which the increase in vaping prevalence has been observed among smokers trying to quit and among ex-smokers who (i) quit ≥1 year ago, (ii) quit recently and did not use an e-cigarette to do so, and (iii) quit before e-cigarettes became popular. We also explored how changes in vaping among ex-smokers differed according to their sociodemographic characteristics and level of alcohol consumption, and whether their profiles (in terms of their duration of abstinence and sociodemographic, drinking, and vaping characteristics) have changed since disposable e-cigarettes started to become popular.

## Methods

### Pre-registration

The study protocol and analysis plan were pre-registered on Open Science Framework (https://osf.io/87tkw/). We amended our planned analyses of trends in recent uptake of vaping after smoking cessation due to the small sample available for this outcome (see *statistical analysis* section for details).

### Design

We analysed data from the Smoking Toolkit Study, a representative repeat cross-sectional survey of adults in England.^13^ The survey began in November 2006 and is ongoing. Each month, a new sample of approximately 1,700 adults is selected via a hybrid of random probability and simple quota sampling. Data were collected face-to-face up to the start of the Covid-19 pandemic and have been collected via telephone interviews since April 2020; the two modalities show good comparability on key sociodemographic and nicotine use indices.^14^

The present analyses focused on data from ex-smokers surveyed between October 2013 (the first wave to assess vaping status among ≥1y ex-smokers) and May 2024 (the most recent data at the time of analysis).

Detailed questions on vaping (beyond current use and use in quit attempts) were included in the survey from July 2016, so we restricted the sample to those surveyed between July 2016 and May 2024 for analyses addressing RQ3 (changes in the profile of ex-smokers who vape).

Vaping characteristics were not assessed in certain waves during this period (May, June, August, September, November, and December 2022; February, March, May, August, September, November, and December 2023; and February, March, and May 2024), so analyses of these variables were limited to those surveyed in eligible waves.

### Measures

Full details of the measures are provided in the study protocol (https://osf.io/87tkw/).

### Smoking status

Smoking status was assessed by asking participants which of the following best applied to them: (a) I smoke cigarettes (including hand-rolled) every day; (b) I smoke cigarettes (including hand-rolled), but not every day; (c) I do not smoke cigarettes at all, but I do smoke tobacco of some kind (e.g., pipe, cigar or shisha); (d) I have stopped smoking completely in the last year; (e) I stopped smoking completely more than a year ago; or (f) I have never been a smoker (i.e., smoked for a year or more). Those who responded *a-d* were considered past-year smokers. Those who responded *d* were considered <1y ex-smokers and those who responded *e* were considered ≥1y ex-smokers.

### Main outcomes

Use of e-cigarettes in quit attempts was assessed among past-year smokers who made at least one attempt to stop smoking in the past year. Quit attempts were assessed with the question: ‘How many serious attempts to stop smoking have you made in the last 12 months? By serious attempt I mean you decided that you would try to make sure you never smoked again. Please include any attempt that you are currently making and please include any successful attempt made within the last year’. Those who reported having made at least one quit attempt were then asked: ‘What did you use to help you stop smoking during the most recent serious quit attempt?’ Those who responded ‘electronic cigarette’ were considered to have used an e-cigarette to support their quit attempt.

Current vaping among ≥1y ex-smokers was assessed with the question: ‘Can I check, are you using any of the following?’. Those who reported using an e-cigarette were considered current vapers.

Early uptake of vaping after smoking cessation was defined as current vaping among <1y ex-smokers who did not use e-cigarettes in their most recent quit attempt,^9^ assessed with the question: ‘Can I check, are you using any of the following either to help you stop smoking, to help you cut down or for any other reason at all?’. Those who reported using an e-cigarette were considered current vapers.

Late uptake of vaping after smoking cessation was defined as current vaping among people who quit smoking before e-cigarettes became popular in 2011.^9^ Duration of abstinence (i.e., how many years ago a participant quit smoking) was calculated as the participant’s actual age minus the age when they stopped smoking. We identified those who quit smoking before 2011 from the year in which they were surveyed and duration of abstinence (e.g., participants surveyed in 2013 with at least 3 years of abstinence, 2014 with at least 4 years of abstinence, 2015 with at least 5 years of abstinence, etc.). Current vaping was assessed as described above, with the question: ‘Can I check, are you using any of the following?’.

### Participant characteristics

Sociodemographic characteristics included age, gender, and occupational social grade (ABC1 includes managerial, professional, and upper supervisory occupations, C2DE includes manual routine, semi-routine, lower supervisory, state pension, and long-term unemployed).

Past-6-month alcohol consumption was assessed with the three-item AUDIT-C. Scores range from 0–12, with higher scores indicating higher levels of consumption. A score of 0 indicates that the participant is a non-drinker, ≤4 is considered low-risk, ≥5 increasing and higher risk, and ≥11 possible dependence. Data on alcohol consumption were only available from March 2014, so analyses by alcohol consumption were limited to this period.

Vaping characteristics included vaping frequency, duration, main device type, usual nicotine strength, and usual source of purchase.

### Statistical analysis

Data were analysed using R v.4.2.1. The Smoking Toolkit Study uses raking to weight the sample to match the population in England.^13^ The following analyses used weighted data. We excluded participants with missing data on smoking or vaping status. Missing cases on other variables were excluded on a per-analysis basis.

### Overall trends in vaping prevalence and uptake among ex-smokers

We used logistic regression to estimate trends across the study period in (i) use of e-cigarettes in attempts to stop smoking, (ii) current vaping among ≥1y ex-smokers, (iii) recent uptake of vaping after smoking cessation among <1y ex-smokers who did not use e-cigarettes in their quit attempt, and (iv) late uptake of vaping after smoking cessation among people who quit smoking before 2011.

Time was modelled using restricted cubic splines, to allow for flexible and non-linear trends. For outcomes (i), (ii), and (iv), we modelled trends by survey month (splines with five knots). We had intended to do the same for recent uptake of vaping after smoking cessation, but sample sizes in each monthly survey wave were too small (mean [SD] monthly number of <1y ex-smokers who did not use e-cigarettes to quit = 14.0 [6.3]; mean [SD] number who vaped = 1.1 [1.2]). We therefore aggregated data annually (12-month periods from October to the following September; e.g., 2013/14 = October 2013 to September 2014, etc.) for this outcome and reduced the number of knots to three so as not to overfit the modelled trend to the reduced number of datapoints. We used predicted estimates from the models to plot trends across the study period.

In a planned sensitivity analysis, we repeated the model for late uptake of vaping after smoking cessation with a restricted sample. We included only those with ≥14 years of abstinence (the minimum duration of abstinence for people who quit before 2011 and who were surveyed in 2024), to reduce the impact of this cohort’s duration of abstinence increasing across the study period (i.e., from ≥3 years for those surveyed in 2013 to ≥14 years for those surveyed in 2024).

### Trends in vaping prevalence and uptake among subgroups of ex-smokers

To explore moderation of trends in vaping among (i) ≥1y ex-smokers and (ii) people who quit smoking before 2011 by age, gender, occupational social grade, and level of alcohol consumption, we repeated each model including the interaction between the moderator of interest and time – thus allowing for time trends to differ across subgroups. Each of the interactions was tested in a separate model. We did not model subgroup trends in recent uptake of vaping after smoking cessation (as planned) because of the small sample size.

Age and alcohol consumption (AUDIT-C) were modelled using restricted cubic splines with three knots (placed at the 5, 50, and 95% percentiles), to allow for non-linear relationships. We displayed estimates for specific ages (18-, 25-, 35-, 45-, 55-, and 65-year-olds) and AUDIT-C scores (0, 3, 6, 9, and 12) to illustrate how trends differ across ages and levels of alcohol consumption. Note that the models used to derive these estimates included data from participants of all ages and AUDIT-C scores.

### Changes in the profile of ex-smokers who vape

We used descriptive statistics to compare the profiles of ≥1y ex-smokers who vaped, before and after disposable e-cigarettes started to become popular in England. Given vaping characteristics were not assessed before July 2016, we restricted this analysis to participants from this wave onwards. In line with evidence showing the rise in use of disposables started around June 2021,^15^ we considered July 2016 to May 2021 to be the pre-disposables period and June 2021 to May 2024 to be the disposables period.^16^

We reported data on quitting history (i.e., duration of abstinence), sociodemographic characteristics, alcohol consumption, and vaping characteristics. We calculated absolute percentage point changes (with 95%CIs) in the proportion belonging to each subgroup (*avg_comparisons* function, *marginaleffects* package^17^). In planned sensitivity analyses, we restricted the pre-disposables period to April-2020 to May-2021 (when data were consistently collected via telephone). Sample sizes were too small to repeat these analyses for ex-smokers who took up vaping after smoking cessation (recent uptake *n*=115 [41/74 pre-disposables/disposables period]; late uptake *n*=196 [59/137]). In an unplanned analysis, we explored changes in mean duration of abstinence among ≥1y ex-smokers who vaped in more detail, aggregating data annually across the entire study period (in 12-month periods from October to November the subsequent year, from October 2013 to May 2024) and modelling the trend using restricted cubic splines (three knots).

## Results

A total of 208,640 adults (≥18y) in England were surveyed between October 2013 and May 2024. We analysed data from 54,251 participants who reported having tried to stop smoking in the past year or having stopped smoking more than a year ago (weighted mean [SD] age = 49.2 [18.2] y; 46.9% women). Characteristics of the subsamples used for each analysis are provided in **Table S1**.

### Trends in vaping prevalence and uptake

Across the study period, there were non-linear changes in the use of e-cigarettes in attempts to stop smoking, current vaping among ≥1y ex-smokers, and recent and late uptake of vaping after smoking cessation (**Figure 1**).

**Figure 1.**
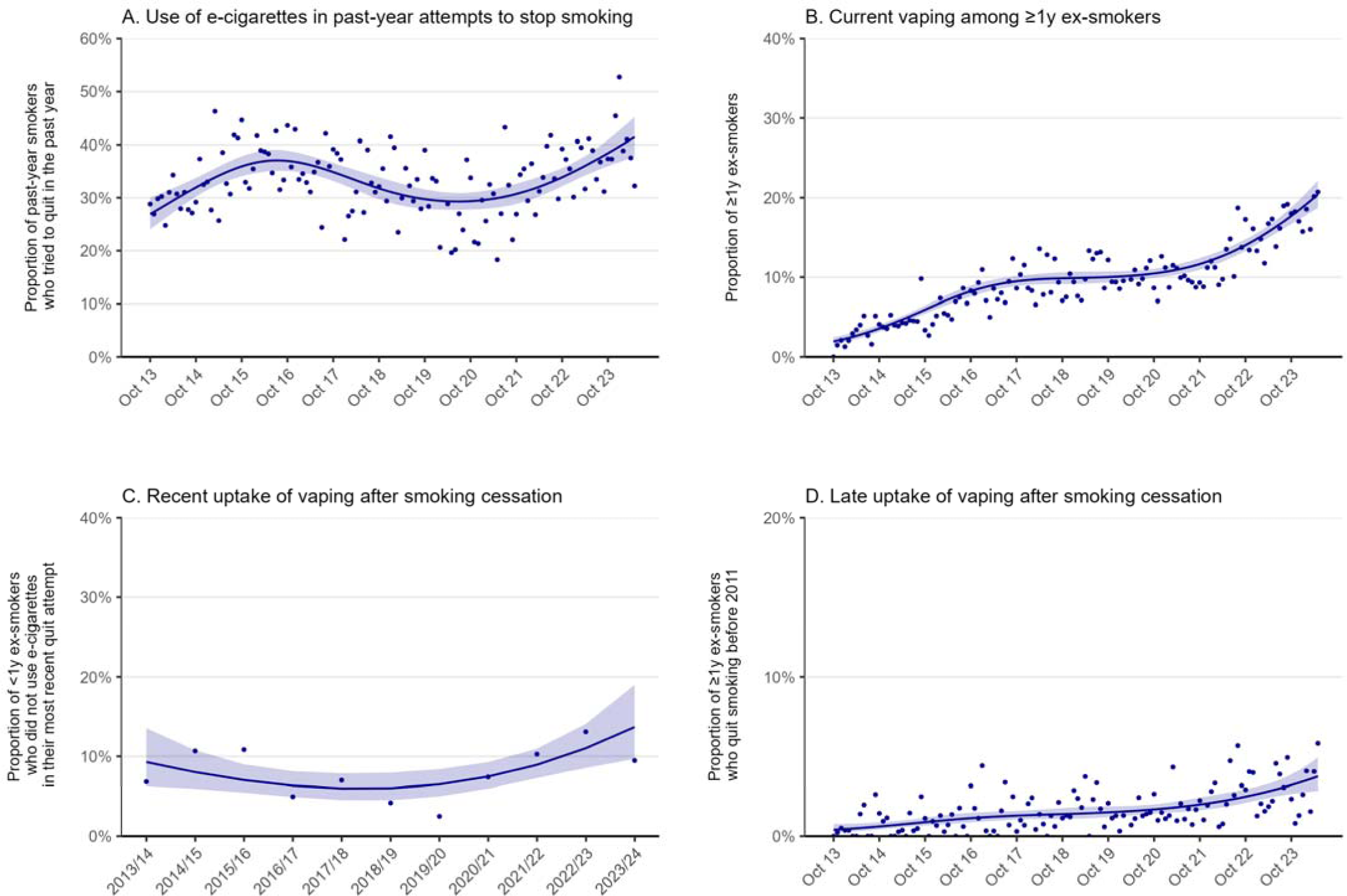
Trends in the use of e-cigarettes for stopping smoking, current vaping among ex-smokers, and uptake of vaping after smoking cessation, October 2013 to May 2024. Panels show the prevalence of (A) e-cigarette use in quit attempts by past-year smokers (*n*=12,593); (B) current vaping among ≥1y ex-smokers (*n*=41,658); (C) current vaping among <1y ex-smokers who did not use e-cigarettes in their most recent quit attempt (*n*=1,782); and (D) current vaping among ≥1y ex-smokers who quit smoking before e-cigarettes started to become popular in 2011 (*n*=29,029). Trends in (A), (B), and (D) were modelled monthly (restricted cubic splines; five knots); (C) was modelled annually (three knots) on account of small samples. Lines represent modelled weighted proportions. Shaded bands represent 95% confidence intervals. Points represent unmodelled weighted proportions.

### Use of e-cigarettes in attempts to stop smoking

Among past-year smokers who tried to quit (*n*=12,593), the proportion who reported using e-cigarettes to support their most recent quit attempt increased from 26.9% [24.0–30.0%] in October 2013 to 37.1% [35.1–39.1%] in July 2016. It then declined to 30.0% [28.1–31.9%] by August 2019 and remained stable for a short period (at an average of 29.6% [27.9–31.3%] between August 2019 and May 2021), before increasing from 30.2% [28.4–32.1%] to a new high of 41.4% [37.7–45.2%] between June 2021 and May 2024 (**Figure 1A**).

### Current vaping among ≥1y ex-smokers

Among ≥1y ex-smokers (*n*=41,658), the proportion who reported current vaping increased from 1.9% [1.5–2.5%] in October 2013 to 9.6% [9.0–10.2%] in December 2017, was relatively stable between December 2017 and May 2021 (at an average of 10.1% [9.5–10.8%]), then increased further from 11.2% [10.6–11.9%] to 20.4% [18.7–22.2%] between June 2021 and May 2024 (**Figure 1B**).

The increase in current vaping among ≥1y ex-smokers was greater at younger ages (e.g., reaching 58.9% among 18-year-olds vs. 10.7% among 65-year-olds; **Figure 2A**, **Table S2**) and among those with the highest levels of alcohol consumption (e.g., reaching 35.4% among those with an AUDIT-C score of 12; **Figure 2D**, **Table S2**). The proportion who vaped was consistently slightly higher among those from less compared with more advantaged social grades, but changes over time were similar (**Figure 2C**). There were no notable differences by gender (**Figure 2B**).

**Figure 2.**
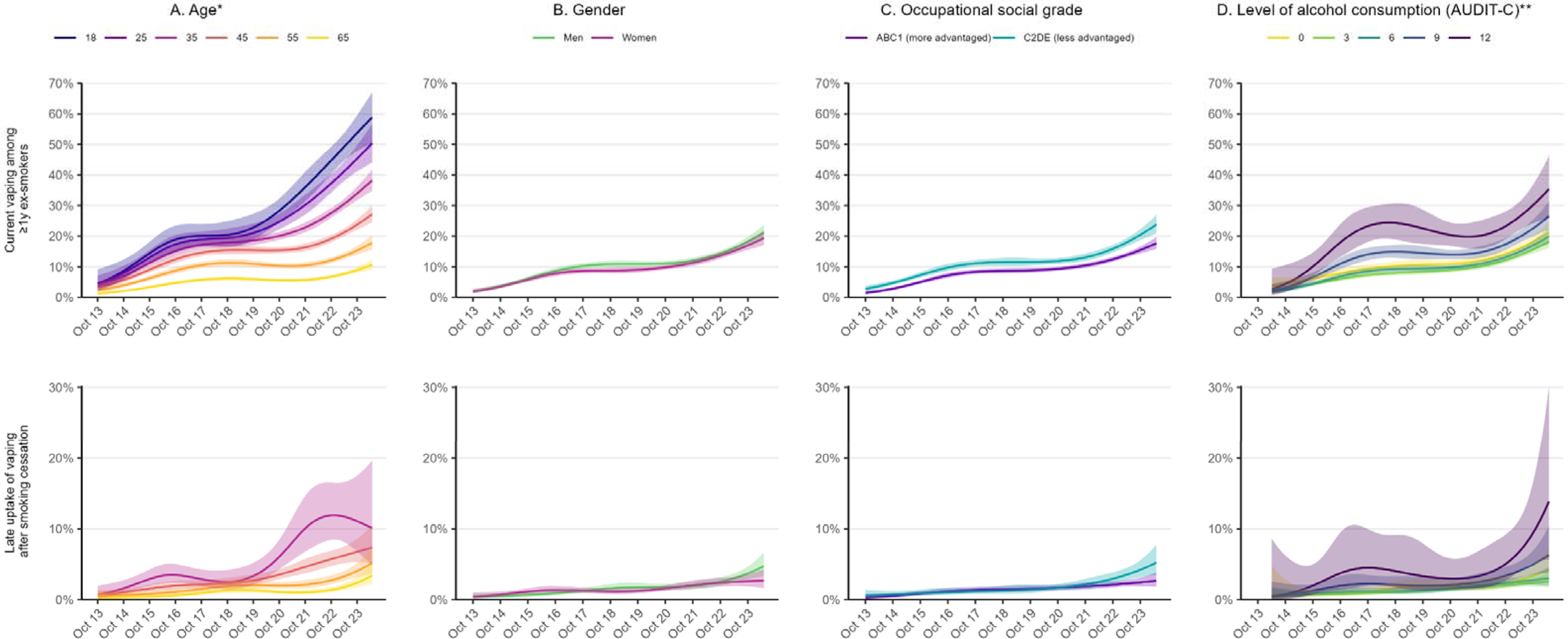
Trends in current vaping and late uptake of vaping after smoking cessation among subgroups of ex-smokers, October 2013 to May 2024. Panels show the prevalence of (i) current vaping among ≥1y ex-smokers and (ii) current vaping among ≥1y ex-smokers who quit smoking before e-cigarettes started to become popular in 2011 (i.e., late uptake of vaping after smoking cessation), by (A) age, (B) gender, (C) occupational social grade, and (D) level of alcohol consumption. Lines represent modelled weighted proportions (time modelled monthly using restricted cubic splines; five knots). Shaded bands represent 95% confidence intervals. *Estimates of late uptake of vaping after smoking cessation are not reported for 18- and 25-year-olds because very few participants in this age range could have quit smoking as an adult before 2011. **Alcohol consumption was assessed from March 2014 onwards. Estimates of prevalence in the first and last months of the time series are provided in **Table S2**.

### Recent uptake of vaping after smoking cessation

Among <1y ex-smokers who reported not using e-cigarettes in their most recent quit attempt (*n*=1,782), there was an uncertain decline in the proportion who reported current vaping between 2013/14 and 2017/18, from 9.3% [6.3–13.5%] to 6.0% [4.5–7.9%], followed by an uncertain increase to 13.7% [9.7–19.0%] between 2017/18 and 2023/24 (**Figure 1C**).

### Late uptake of vaping after smoking cessation

Among ≥1y ex-smokers who quit smoking before e-cigarettes started to become popular in 2011 (*n*=29,029), the proportion who reported current vaping increased from 0.4% [0.2–0.8%] in October 2013 to 1.9% [1.6–2.3%] in June 2021 then increased more rapidly, reaching 3.7% [2.8–4.9%] by May 2024 (**Figure 1D**). The trend was similar when we restricted this group to

≥14y ex-smokers (to ensure a more consistent duration of abstinence within this group across the period), although absolute prevalence estimates were lower in earlier months: increasing from 0.01% [0.0–0.3%] in October 2013 to 1.1% [0.8–1.4%] in June 2021 and to 3.6% [2.6–5.0%] in May 2024 (**Figure S1A**).

The increase in current vaping among ≥1y ex-smokers who quit smoking before e-cigarettes started to become popular in 2011 appeared more pronounced among those who were younger (e.g., reaching 10.1% among 35-year-olds vs. 3.4% among 65-year-olds; **Figure 2A**) and those who drank more heavily (e.g., reaching 13.9% among those with an AUDIT-C score of 12; **Figure 2D**), although 95% CIs were wide, introducing some uncertainty (**Table S2**). There were also potentially smaller differences by gender and occupational social grade near the end of the period, with rates rising to higher levels among men than women (4.7% vs. 2.7%; **Figure 2B**) and those from less vs. more advantaged social grades (5.2% vs. 2.7%; **Figure 2C**), but these differences were uncertain (**Table S2**).

### Changes in the profile of ex-smokers who vape

There were several changes in the profile of ≥1y ex-smokers who vaped from before (July 2016 to May 2021) to after (June 2021 to May 2024) disposable e-cigarettes started to become popular (**Table 1**).

**Table 1.**
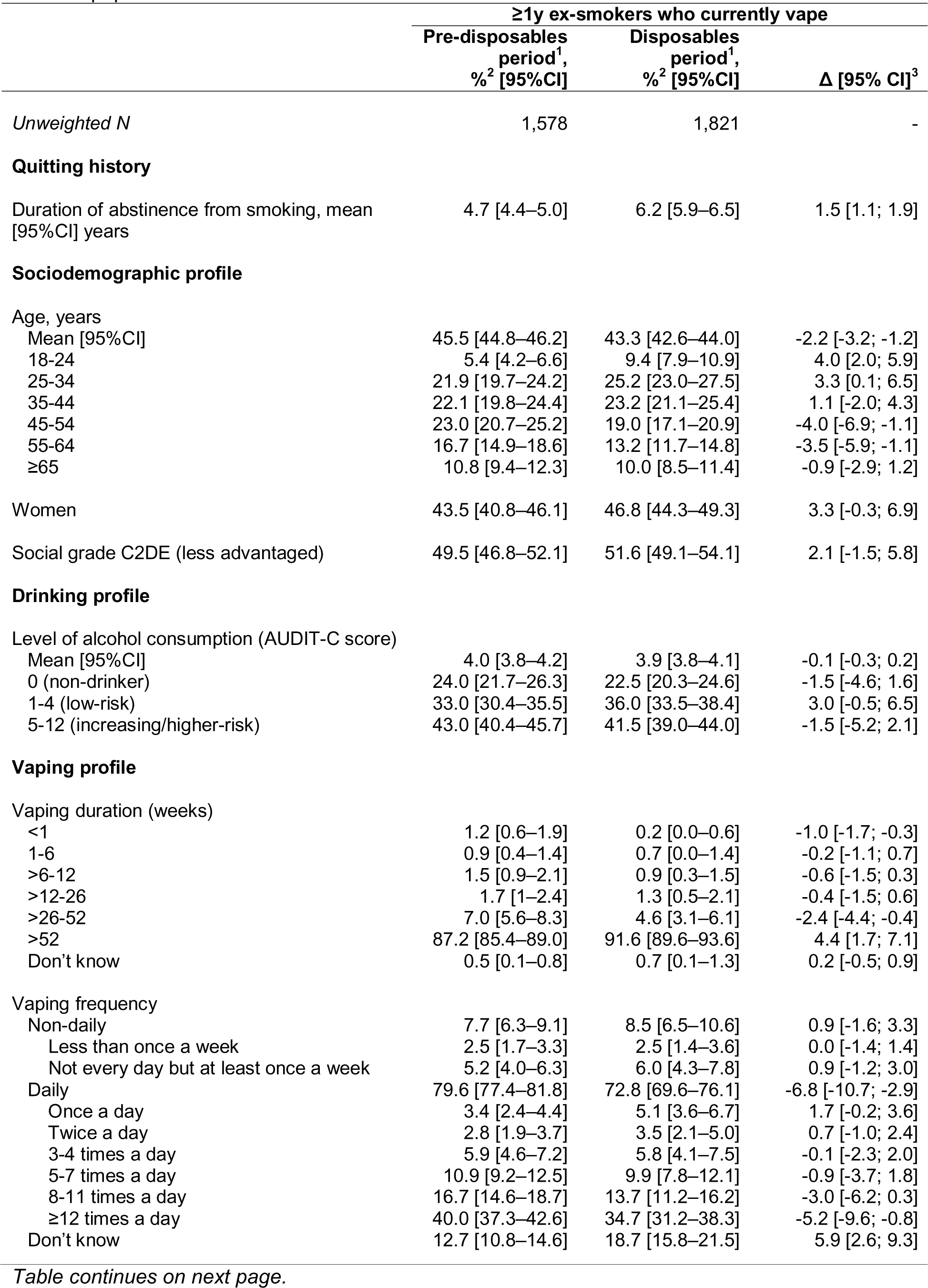
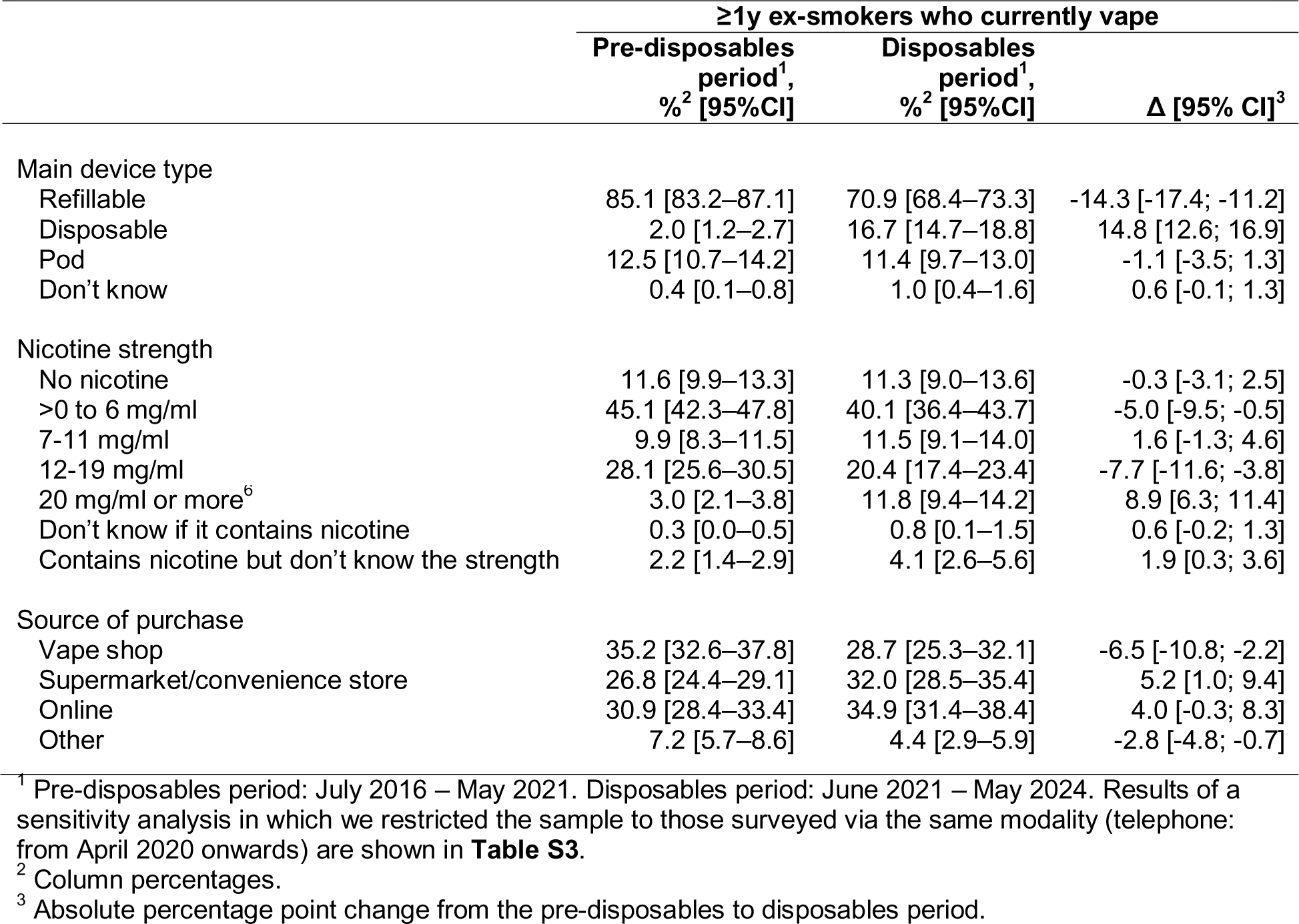
Changes in the profile of ex-smokers who vape since disposable e-cigarettes started to become popular.

Ex-smoking vapers surveyed since disposables started to become popular reported a greater duration of abstinence from smoking, on average, than those surveyed earlier (6.2 vs. 4.7y). Unplanned analyses showed the mean duration of abstinence among ex-smoking vapers increased non-linearly across the entire study period, with modelled estimates increasing from 2.1y [1.6–2.5y] in 2013/14 to 5.6y [5.3–5.9y] in 2019/20 and then appearing to level off (reaching 6.2y [5.8–6.7y] in 2023/24; **Figure S2**). In addition, a greater proportion were aged 18-34y (34.6% vs. 27.3%). There were no notable changes in terms of gender, occupational social grade, or alcohol consumption.

In terms of vaping characteristics, ex-smoking vapers surveyed since disposables started to become popular were more likely than those surveyed earlier to report having been vaping for more than a year (91.6% vs. 87.2%) and less likely to report having been vaping for <1 week (0.2% vs. 1.2%) or for between 6 months and a year (4.6% vs. 7.0%). They were also more likely to say they did not know how frequently they vaped (18.7% vs. 12.7%) and less likely to report vaping daily (72.8% vs. 79.6%). They were much more likely to report mainly using disposable e-cigarettes (16.7% vs. 2.0%), offset by a decline in the use of refillable devices (70.9% vs. 85.1%). They were more likely to report using the highest-strength (≥20 mg/ml) nicotine e-liquids (11.8% vs. 3.0%) or to say that they did not know the strength of their nicotine-containing e-liquid (4.1% vs. 2.2%). They were also more likely to report buying their vaping products from supermarkets and convenience stores (32.0% vs. 26.8%) and less likely to report buying them from vape shops (28.7% vs. 35.2%) or ‘other’ sources (4.4% vs. 7.2%).

A sensitivity analysis restricted to participants who were interviewed via telephone (April 2020 onwards; **Table S3**) showed a broadly similar pattern. However, there were two notable differences. First, when the pre-disposables period was shortened (starting at April 2020 rather than July 2016), vaping duration was more similar across ex-smoking vapers surveyed after compared with before disposables started to become popular (e.g., 91.6% vs. 93.5% reported having vaped for more than a year). Second, in addition to an increase in the proportion of ex-smoking vapers who reported mainly buying their products from supermarkets and convenience stores since disposables started to become popular (32.0% vs. 26.5%), the proportion who reported mainly buying from vape shops increased (28.7% vs. 23.4%) and the proportion mainly buying from online sources decreased (34.9% vs. 42.6%).

## Discussion

Over the past decade, there have been clear shifts in vaping prevalence and uptake among adults in England who have quit smoking. In October 2013, when e-cigarettes were still fairly new and delivered nicotine less efficiently, around one in 50 ≥1y ex-smokers vaped. This number increased steadily to one in ten by the end of 2017 and remained stable for several years. It then increased sharply from 2021, reaching one in five by May 2024, equivalent to approximately 2.2 million people (45.2 million adults ≥18y in England^18^ * 23.8% ≥1y ex-smokers [Smoking Toolkit Study, January–May 2024] * 20.4% vaping prevalence). This pattern is consistent with that observed in the general adult population: an initial rise in popularity of e-cigarettes, followed by a plateau and then a subsequent rapid rise linked to the introduction of new disposable e-cigarettes to the market.^10^

Much of this increase in vaping prevalence among ex-smokers is likely to be the result of more people using e-cigarettes as a smoking cessation aid who continue to use them after stopping smoking. The timing of the changes we observed coincided with changes in the use of e-cigarettes by people attempting to quit smoking. Studies have shown that a substantial proportion of those who quit with the support of an e-cigarette continue to vape for many months (and in some cases, years) beyond their successful quit attempt. For example, a randomised controlled trial of e-cigarettes vs. nicotine replacement therapy for smoking cessation found that among those in the e-cigarette condition who were abstinent at one year, 80% were still vaping.^19^ UK guidance advises people not to rush to stop vaping after quitting smoking, but rather to gradually reduce their vaping frequency or nicotine strength when they feel confident that they can do this without going back to smoking.^20^ As such, one would expect to see an increase in vaping among ex-smokers as use of e-cigarettes in quit attempts increases.

However, not all of the increase in vaping among ex-smokers was attributable to continued vaping after successfully quitting smoking with an e-cigarette. Our data also provide evidence of an increase in the uptake of vaping *after* successful smoking cessation. In October 2013, vaping was rare among people who quit smoking before e-cigarettes started to become popular in 2011, at around one in 250 ex-smokers. By May 2024, this number had increased to one in 27, equivalent to approximately 212,000 people (45.2 million adults ≥18y in England^18^ * 12.7% ex-smokers who quit before 2011 [Smoking Toolkit Study, January–May 2024] * 3.7% vaping prevalence). When we set the minimum duration of abstinence constant across this period (at ≥14y) the increase was even more stark, with the number of vapers increasing from one in 10,000 to one in 27. Changes in the uptake of vaping among recent (<1y) ex-smokers were uncertain, at least partially due to smaller sample sizes, but also suggested a possible increase in recent years.

Uptake of vaping among ex-smokers may be influenced by new product developments and social trends. Increases in vaping prevalence were greatest at younger ages, among whom disposable e-cigarettes (and as a result, vaping more generally) have become particularly popular since 2021.^11,15^ These influences may be greater among those with a propensity for risk-taking behaviour.^21^ Consistent with this, increases in vaping among ex-smokers were also larger among those who reported the highest levels of alcohol consumption. A similar pattern has been documented among adults who have never regularly smoked.^12^

In terms of the profile of ex-smokers who vape, we observed several changes since disposable e-cigarettes started to become popular. Most of these reflect changes that have occurred among vapers more generally: younger age, increased use of disposables and higher nicotine strengths, and increased purchasing from supermarkets and convenience stores.^11,12,22,23^ Ex-smoking vapers surveyed more recently also reported a longer duration of abstinence from smoking, on average, than those surveyed earlier. This may partly reflect ex-smokers who vape accumulating over time as people take up vaping and continue to vape long-term (echoed by results indicating more are vaping for >1 year). It may also reflect increased uptake of vaping among long-term ex-smokers or more recent ex-smokers relapsing back to smoking.

The health impacts of people taking up vaping after having stopped smoking will depend on what they would be doing if they did not vape. If they would otherwise not use nicotine, there is a risk that starting to vape may increase their risk of relapse to smoking by reintroducing them to regular nicotine exposure (although people typically report lower levels of dependence on vaping than smoking^24^). Vaping, while much less harmful than smoking, will also expose long-term ex-smokers to more harm than not vaping or smoking.^5^ However, if ex-smokers take up vaping instead of relapsing to smoking this will reduce the harm they are exposed to.^5^ Among very long-term ex-smokers, the risk of relapse would be low,^25^ so taking up vaping is probably more likely to have unintended consequences (i.e., exposure to harm, increased risk of relapse) than benefits. More research is needed to better understand the extent to which vaping increases vs. reduces the risk of relapse to smoking (both among ex-smokers who vape continuously from the point of a successful quit attempt and among those who take up vaping after quitting smoking) in different tobacco and nicotine regulatory contexts and markets. As with examining whether e-cigarettes act as a causal gateway to smoking among youth,^26^ this research should triangulate evidence from both the individual- and population-level using diverse methodologies with different sources of bias, and in priority groups that exhibit differential risks of returning to smoking.

The plateau in current vaping among long-term ex-smokers between 2018 and 2021 has a number of possible explanations. If people were quitting smoking with the use of e-cigarettes at a broadly constant rate, and continuing to vape long-term (with a proportion eventually quitting vaping too) at similar rates, then one would expect the proportion of long-term ex-smokers who were vaping to grow at a broadly linear rate, providing the rates of vaping uptake after cessation were also constant (which we observed during this period in the current study). Thus, the observed plateau between 2018 and 2021 (going against the previously observed steady increase) may reflect an increase during that period of long-term ex-smokers quitting vaping and/or relapsing to smoking. During that period, we also saw the average duration of smoking abstinence among long-term ex-smokers who vaped increase up to around 2019 but there appeared to be a plateau thereafter. In separate studies, we have observed a slowing in overall smoking prevalence around the same period,^27^ as well as increases in the proportion of non-daily smokers^28^ and increases in the prevalence of the dual use of e-cigarettes and cigarettes.^29^ All of which is consistent with increased relapse to non-daily smoking among long-term ex-smokers from around 2018 onwards. Insofar that this occurred, the cause(s) is unclear but coincided with big increases in the risk perceptions of e-cigarettes^30^ and the onset of the covid pandemic and its associated impacts. When formulating vaping policy, any effects on relapse to long-term ex-smokers who vape may represent a serious and unintended public health risk to be considered for countries in which large numbers of people have already switched from smoking to vaping.

Strengths of this study include the representative sample, monthly data collection, and comprehensive assessment of vaping behaviour. There were also limitations. The reliance on self-reported data may introduce recall bias, particularly for events that happened a long time ago (e.g., how long ago the participant quit smoking). In addition, while the sample was large overall, small sample sizes for certain subgroups (e.g., recent ex-smokers) limited the precision of some estimates. Findings cannot be presumed to generalise to other countries.

In conclusion, vaping prevalence increased substantially among adult ex-smokers in England over the past decade, particularly at younger ages. While this is likely to have been partly driven by increases in people using e-cigarettes as a smoking cessation aid and continuing to vape beyond their successful quit attempt, there was also evidence of increased uptake of vaping among those who had been abstinent from smoking for many years.

## Data Availability

Data are available upon reasonable request to the authors.

## Declarations

### Data sharing

Data are available upon reasonable request to the authors.

### Ethics approval

Ethical approval for the STS was granted originally by the UCL Ethics Committee (ID 0498/001). The data are not collected by UCL and are anonymised when received by UCL.

### Competing interests

JB has received unrestricted research funding from Pfizer and J&J, who manufacture smoking cessation medications. LS has received honoraria for talks, unrestricted research grants and travel expenses to attend meetings and workshops from manufactures of smoking cessation medications (Pfizer; J&J), and has acted as paid reviewer for grant awarding bodies and as a paid consultant for health care companies. All authors declare no financial links with tobacco companies, e-cigarette manufacturers, or their representatives.

### Funding

This work was supported by Cancer Research UK (PRCRPG-Nov21\100002). For the purpose of Open Access, the author has applied a CC BY public copyright licence to any Author Accepted Manuscript version arising from this submission.

## Notes

### Clinical Protocols

https://osf.io/87tkw/

