## Supplementary material for "Prevalence and uptake of vaping among people who have quit smoking: a population study in England, 2013-2024": Table S1

### Table S1. Sample characteristics

|  | **Whole sample** | **Past-year smokers** | **≥1y ex-smokers** | **<1y ex-smokers who did not use e-cigarettes in their quit attempt** | **≥1y ex-smokers who quit before 2011** |
| --- | --- | --- | --- | --- | --- |
| Unweighted *N* | 80,472 | 12,593 | 41,658 | 1,782 | 29,029 |
| Age (years) |  |  |  |  |  |
| Mean (SD) | 49.2 (18.2) | 39.4 (15.4) | 56.4 (17.0) | 40.1 (15.8) | 62.1 (14.6) |
| 18-24 | 9.8% | 19.2% | 2.8% | 18.8% | 0.2% |
| 25-34 | 17.1% | 27.5% | 10.1% | 27.0% | 3.1% |
| 35-44 | 16.2% | 19.0% | 14.4% | 19.5% | 10.9% |
| 45-54 | 17.6% | 15.9% | 17.9% | 15.0% | 17.6% |
| 55-64 | 15.2% | 10.5% | 18.0% | 10.2% | 20.2% |
| ≥65 | 24.1% | 7.9% | 36.7% | 9.5% | 48.1% |
| Gender |  |  |  |  |  |
| Men | 52.8% | 51.8% | 52.3% | 52.0% | 52.6% |
| Women | 46.9% | 47.7% | 47.5% | 47.5% | 47.3% |
| Other | 0.3% | 0.5% | 0.2% | 0.5% | 0.1% |
| *Missing* | 105 | 22 | 41 | 2 | 13 |
| Occupational social grade |  |  |  |  |  |
| ABC1 (more advantaged) | 48.9% | 42.5% | 57.1% | 51.7% | 59.6% |
| C2DE (less advantaged) | 51.1% | 57.5% | 42.9% | 48.3% | 40.4% |
| Alcohol consumption^1^ |  |  |  |  |  |
| Mean (SD) AUDIT-C score | 3.7 (3.2) | 3.6 (3.3) | 3.7 (2.9) | 3.6 (3.2) | 3.6 (2.9) |
| 0 (non-drinker) | 24.6% | 30.7% | 19.4% | 27.8% | 17.9% |
| 1-4 (low-risk) | 38.0% | 31.5% | 44.7% | 34.4% | 47.6% |
| 5-12 (increasing/higher-risk) | 37.4% | 37.7% | 35.8% | 37.8% | 34.5% |
| *Missing* | 4,821 | 920 | 2,165 | 128 | 1,756 |

^1^ Alcohol consumption was assessed from April 2014 onwards. The number of missing cases includes those surveyed between October 2013 and March 2014.

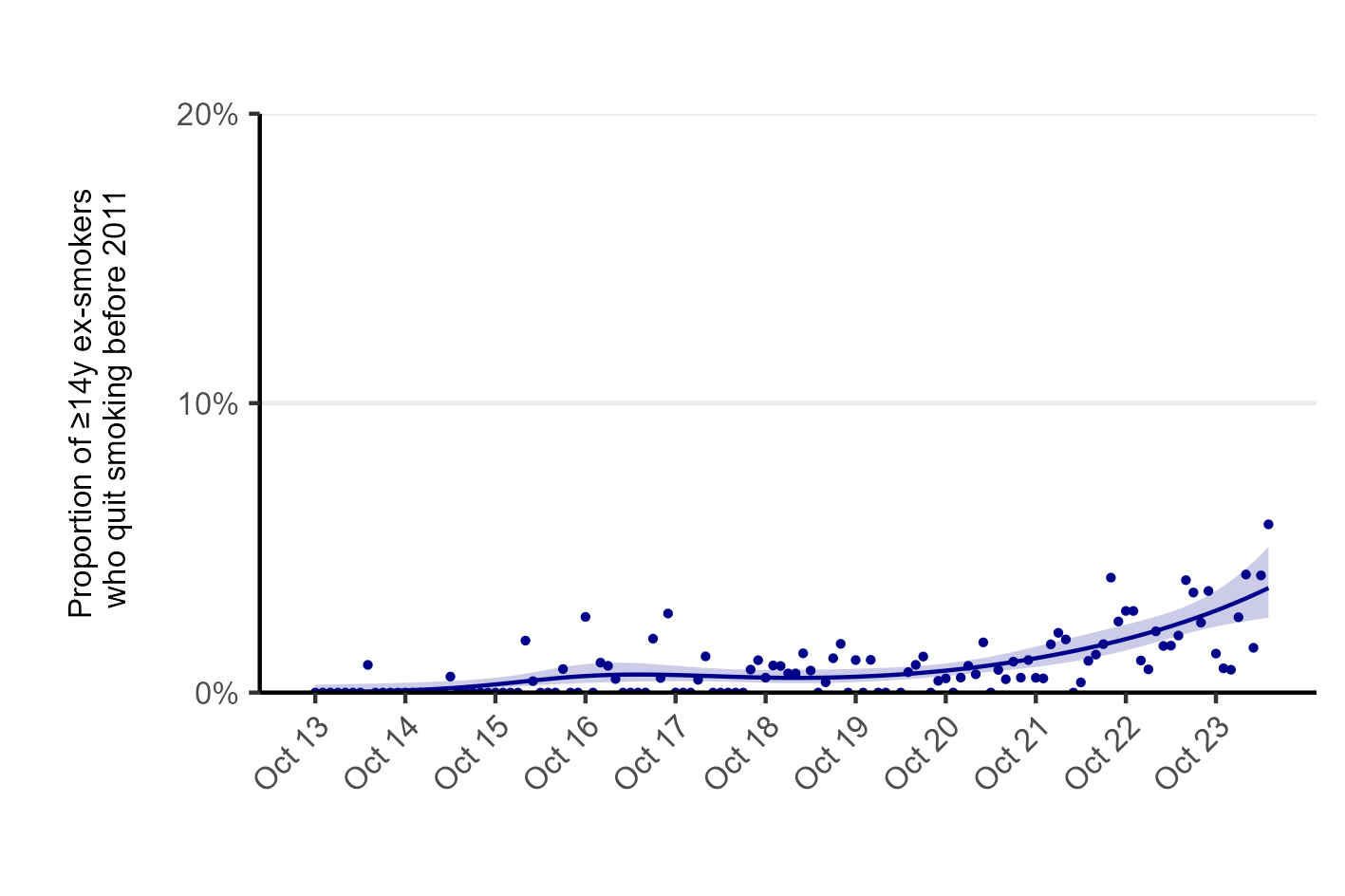

### Figure S1. Trend in late uptake of vaping after smoking cessation (≥14y ex-smokers), October 2013 to May 2024. Prevalence of current vaping among ≥14y ex-smokers who quit smoking before e-cigarettes started to become popular in 2011 (*n*=23,740). Line represents the modelled weighted proportion by monthly survey wave (modelled non-linearly using restricted cubic splines with five knots). Shaded band represents 95% confidence intervals. Points represent the unmodelled weighted proportion by month.

### Table S2. Modelled estimates of the prevalence of current vaping and late uptake of vaping after smoking cessation among subgroups of ex-smokers in England, in the first and last months of the study period

|  | **Prevalence, % [95%CI]^1^** | | | | |
| --- | --- | --- | --- | --- | --- |
|  | **Current vaping^2^** | |  | **Late uptake after smoking cessation^3^** | |
|  | **Oct 2013** | **May 2024** |  | **Oct 2013** | **May 2024** |
| Year of age^4^ |  |  |  |  |  |
| 18 | 4.5 [2.1–9.2] | 58.9 [50.3–67.0] |  | - | - |
| 25 | 4.2 [2.5–7.1] | 50.4 [44.1–56.7] |  | - | - |
| 35 | 3.9 [2.8–5.2] | 38.3 [34.8–41.8] |  | 0.7 [0.3–2.0] | 10.1 [4.9–19.7] |
| 45 | 3.3 [2.4–4.4] | 27.2 [24.5–30.0] |  | 0.6 [0.3–1.3] | 7.4 [5.2–10.5] |
| 55 | 2.3 [1.6–3.3] | 17.8 [15.6–20.4] |  | 0.5 [0.2–1.3] | 5.2 [3.6–7.4] |
| 65 | 1.2 [0.8–1.8] | 10.7 [9.1–12.5] |  | 0.3 [0.1–0.8] | 3.4 [2.2–5.3] |
| Gender |  |  |  |  |  |
| Men | 2.0 [1.4–2.7] | 21.2 [18.8–23.7] |  | 0.4 [0.2–1.0] | 4.7 [3.3–6.7] |
| Women | 1.9 [1.4–2.7] | 19.3 [17.0–21.9] |  | 0.4 [0.2–1.0] | 2.7 [1.7–4.3] |
| Occupational social grade |  |  |  |  |  |
| ABC1 (more advantaged) | 1.4 [1.0–2.1] | 17.7 [15.8–19.7] |  | 0.3 [0.1–0.7] | 2.7 [1.8–3.8] |
| C2DE (less advantaged) | 2.8 [2.0–3.9] | 23.8 [21.0–27.0] |  | 0.6 [0.2–1.4] | 5.2 [3.5–7.8] |
|  | **Current vaping^2^** | |  | **Late uptake after smoking cessation^3^** | |
|  | **Apr 2014** | **May 2024** |  | **Apr 2014** | **May 2024** |
| Level of alcohol consumption (AUDIT-C score)^5^ |  |  |  |  |  |
| 0 (lowest) | 3.8 [2.2–6.4] | 21.1 [18.0–24.7] |  | 0.7 [0.1–4.8] | 4.2 [2.5–7.2] |
| 3 | 2.2 [1.5–3.2] | 18.2 [16.1–20.5] |  | 0.5 [0.2–1.6] | 2.5 [1.8–3.7] |
| 6 | 1.8 [1.1–3.0] | 19.9 [17.6–22.5] |  | 0.5 [0.1–1.8] | 3.0 [2.0–4.6] |
| 9 | 2.2 [1.1–4.4] | 26.6 [22.2–31.4] |  | 0.5 [0.1–2.6] | 6.3 [3.8–10.4] |
| 12 (highest) | 2.7 [0.8–9.4] | 35.4 [25.9–46.3] |  | 0.5 [0.0–8.6] | 13.9 [5.7–30.1] |

^1^ Data are weighted estimates of prevalence in the first and last months in the study period from logistic regression with survey month modelled non-linearly using restricted cubic splines (five knots).

^2^ Current vaping among ≥1y ex-smokers.

^3^ Current vaping among ex-smokers who quit smoking before e-cigarettes became popular in 2011.

^4^ Modelled estimates for selected ages. Note that the model used to derive these estimates included data from participants of all ages (≥18y), not only those who were aged exactly 18, 25, 35, 45, 55, or 65 years. We do not report estimates of the prevalence of late uptake of vaping after smoking cessation among 18- and 25-year-olds because very few participants in this age range could have quit smoking as an adult before 2011.

^5^ AUDIT-C scores range from 0 to 12. Note that the model used to derive these estimates included data from participants with any score on this scale, not only those with a score of exactly 0, 3, 6, 9, or 12. AUDIT-C data first collected April 2014.

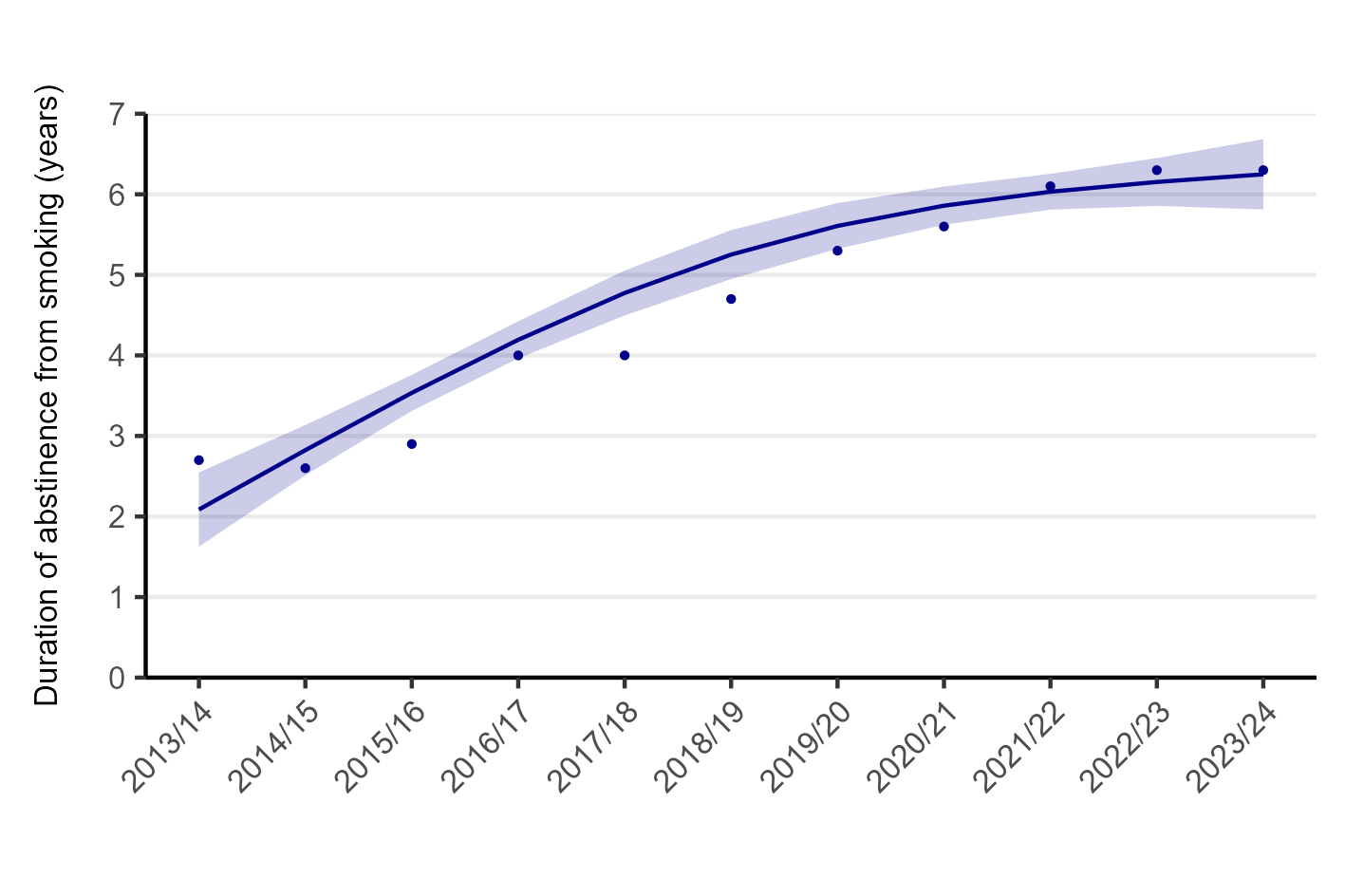

### Figure S2. Trend in the mean duration of abstinence from smoking by survey year among ex-smokers who vape, October 2013 to May 2024. Mean duration of abstinence among ≥1y ex-smokers who vape. Line represents the modelled weighted proportion by survey year (12-month periods from October to the following September [e.g., 2013/14 = October 2013 to September 2014, etc.]; modelled non-linearly using restricted cubic splines with three knots). Shaded band represents 95% confidence intervals. Points represent the unmodelled weighted proportion by year.

### Table S3. Changes in the profile of ex-smokers who vape since disposable e-cigarettes started to become popular

|  | **≥1y ex-smokers who currently vape** | | |
| --- | --- | --- | --- |
|  | **Pre-disposables period^1^,**  **%^2^ [95%CI]** | **Disposables period^1^,**  **%^2^ [95%CI]** | **Δ [95% CI]^3^** |
| *Unweighted N* | 529 | 1,821 | - |
| **Quitting history** |  |  |  |
| Duration of abstinence from smoking, mean (SD) years | 5.3 [5.0–5.7] | 6.2 [5.9–6.5] | 0.9 [0.4; 1.3] |
| **Sociodemographic profile** |  |  |  |
| Age, years |  |  |  |
| Mean (SD) | 45.1 [43.8–46.4] | 43.3 [42.6–44] | -1.8 [-3.3; -0.3] |
| 18-24 | 6.4 [4.0–8.7] | 9.4 [7.9–10.9] | 3.0 [0.2; 5.9] |
| 25-34 | 22.4 [18.5–26.3] | 25.2 [23.0–27.5] | 2.8 [-1.7; 7.3] |
| 35-44 | 23.6 [19.5–27.7] | 23.2 [21.1–25.4] | -0.4 [-5.0; 4.3] |
| 45-54 | 21.2 [17.5–24.8] | 19.0 [17.1–20.9] | -2.2 [-6.3; 1.9] |
| 55-64 | 15.5 [12.4–18.6] | 13.2 [11.7–14.8] | -2.3 [-5.7; 1.2] |
| ≥65 | 11.0 [8.4–13.6] | 10.0 [8.5–11.4] | -1.0 [-4.0; 1.9] |
| Women | 43.6 [39.1–48.1] | 46.8 [44.3–49.3] | 3.2 [-1.9; 8.3] |
| Social grade C2DE (less advantaged) | 49.9 [45.3–54.4] | 51.6 [49.1–54.1] | 1.7 [-3.5; 6.9] |
| **Drinking profile** |  |  |  |
| Level of alcohol consumption (AUDIT-C score) |  |  |  |
| Mean (SD) | 4.0 [3.7–4.3] | 3.9 [3.8–4.1] | -0.1 [-0.4; 0.3] |
| 0 (non-drinker) | 21.9 [18.0–25.8] | 22.5 [20.3–24.6] | 0.6 [-3.9; 5.0] |
| 1-4 (low-risk) | 34.4 [29.9–38.9] | 36.0 [33.5–38.4] | 1.6 [-3.5; 6.7] |
| 5-12 (increasing/higher-risk) | 43.7 [39.1–48.3] | 41.5 [39.0–44.0] | -2.2 [-7.4; 3.1] |
| **Vaping profile** |  |  |  |
| Vaping duration (weeks) |  |  |  |
| <1 | 0 | 0.2 [0.0–0.6] | 0.2 [-0.1; 0.6] |
| 1-6 | 0 | 0.7 [0.0–1.4] | 0.7 [0.0; 1.4] |
| >6-12 | 0.2 [0.0–0.5] | 0.9 [0.3–1.5] | 0.7 [0.1; 1.4] |
| >12-26 | 1.4 [0.3–2.5] | 1.3 [0.5–2.1] | -0.1 [-1.4; 1.2] |
| >26-52 | 4.7 [2.8–6.6] | 4.6 [3.1–6.1] | -0.1 [-2.5; 2.3] |
| >52 | 93.5 [91.3–95.7] | 91.6 [89.6–93.6] | -1.9 [-4.9; 1.1] |
| Don’t know | 0.3 [0.0–0.7] | 0.7 [0.1–1.3] | 0.4 [-0.3; 1.1] |
| Vaping frequency |  |  |  |
| Non-daily | 9.3 [6.6–12.1] | 8.5 [6.5–10.6] | -0.8 [-4.2; 2.6] |
| Less than once a week | 2.4 [1.0–3.7] | 2.5 [1.4–3.6] | 0.1 [-1.6; 1.9] |
| Not every day but at least once a week | 7.0 [4.5–9.4] | 6.0 [4.3–7.8] | -0.9 [-3.9; 2.0] |
| Daily | 73.9 [69.7–78.1] | 72.8 [69.6–76.1] | -1.1 [-6.3; 4.2] |
| Once a day | 3.7 [1.7–5.6] | 5.1 [3.6–6.7] | 1.5 [-1.0; 4.0] |
| Twice a day | 4.1 [2.1–6.2] | 3.5 [2.1–5.0] | -0.6 [-3.1; 1.9] |
| 3-4 times a day | 6.0 [3.7–8.3] | 5.8 [4.1–7.5] | -0.2 [-3.1; 2.7] |
| 5-7 times a day | 9.3 [6.6–12.0] | 9.9 [7.8–12.1] | 0.7 [-2.8; 4.1] |
| 8-11 times a day | 15.5 [12.0–18.9] | 13.7 [11.2–16.2] | -1.8 [-6.0; 2.5] |
| ≥12 times a day | 35.4 [30.8–40.0] | 34.7 [31.2–38.3] | -0.6 [-6.4; 5.1] |
| Don’t know | 16.8 [13.2–20.3] | 18.7 [15.8–21.5] | 1.9 [-2.6; 6.4] |

*Table continues on next page.*

**Table S3.** *continued*

|  | **≥1y ex-smokers who currently vape** | | |
| --- | --- | --- | --- |
|  | **Pre-disposables period^1^,**  **%^2^ [95%CI]** | **Disposables period^1^,**  **%^2^ [95%CI]** | **Δ [95% CI]^3^** |
| Main device type |  |  |  |
| Refillable | 87.7 [84.6–90.7] | 70.9 [68.4–73.3] | -16.8 [-20.7; -12.9] |
| Disposable | 1.0 [0.1–1.8] | 16.7 [14.7–18.8] | 15.8 [13.6; 18.0] |
| Pod | 11.0 [8.0–13.9] | 11.4 [9.7–13.0] | 0.4 [-3.0; 3.8] |
| Don’t know | 0.4 [0.0–0.8] | 1.0 [0.4–1.6] | 0.6 [-0.1; 1.4] |
| Nicotine strength |  |  |  |
| No nicotine | 12.8 [9.6–16.1] | 11.3 [9.0–13.6] | -1.5 [-5.5; 2.4] |
| >0 to 6 mg/ml | 47.7 [42.9–52.6] | 40.1 [36.4–43.7] | -7.7 [-13.7; -1.7] |
| 7-11 mg/ml | 7.5 [5.1–10.0] | 11.5 [9.1–14.0] | 4.0 [0.5; 7.5] |
| 12-19 mg/ml | 25.9 [21.7–30.0] | 20.4 [17.4–23.4] | -5.5 [-10.6; -0.4] |
| 20 mg/ml or more^6^ | 1.6 [0.4–2.7] | 11.8 [9.4–14.2] | 10.2 [7.6; 12.9] |
| Don’t know if it contains nicotine | 0.5 [0.0–1.0] | 0.8 [0.1–1.5] | 0.3 [-0.5; 1.2] |
| Contains nicotine but don’t know the strength | 4.0 [2.2–5.8] | 4.1 [2.6–5.6] | 0.1 [-2.3; 2.4] |
| Source of purchase |  |  |  |
| Vape shop | 23.4 [19.4–27.4] | 28.7 [25.3–32.1] | 5.3 [0.0; 10.5] |
| Supermarket/convenience store | 26.5 [22.3–30.7] | 32.0 [28.5–35.4] | 5.5 [0.0; 10.9] |
| Online | 42.6 [37.8–47.4] | 34.9 [31.4–38.4] | -7.6 [-13.6; -1.7] |
| Other | 7.5 [4.8–10.3] | 4.4 [2.9–5.9] | -3.1 [-6.3; 0.0] |

^1^ Pre-disposables period: Apr 2020 – May 2021. Disposables period: June 2021 – May 2024.

^2^ Column percentages.

^3^ Absolute percentage point change from the pre-disposables to disposables period.
